# RBD amplicon sequencing of wastewater reveals patterns of variant emergence and evolution

**DOI:** 10.1101/2024.07.12.24310301

**Authors:** Xingwen Chen, John Balliew, Cici X. Bauer, Jennifer Deegan, Anna Gitter, Blake M. Hanson, Anthony W. Maresso, Michael J. Tisza, Catherine L. Troisi, Janelle Rios, Kristina D. Mena, Eric Boerwinkle, Fuqing Wu

## Abstract

Rapid evolution of SARS-CoV-2 has resulted in the emergence of numerous variants, posing significant challenges to public health surveillance. Clinical genome sequencing, while valuable, has limitations in capturing the full epidemiological dynamics of circulating variants in the general population. This study utilized receptor-binding domain (RBD) amplicon sequencing of wastewater samples to monitor the SARS-CoV-2 community dynamics and evolution in El Paso, TX. Over 17 months, we identified 91 variants and observed waves of dominant variants transitioning from BA.2 to BA.2.12.1, BA.4&5, BQ.1, and XBB.1.5. Our findings demonstrated early detection of variants and identification of unreported outbreaks, while showing strong consistency with clinical genome sequencing data at the local, state, and national levels. Alpha diversity analyses revealed significant periodical variations, with the highest diversity observed in winter and the outbreak lag phases, likely due to lower competition among variants before the outbreak growth phase. The data underscores the importance of low transmission periods for rapid mutation and variant evolution. This study highlights the effectiveness of integrating RBD amplicon sequencing with wastewater surveillance in tracking viral evolution, understanding variant emergence, and enhancing public health preparedness.

## Introduction

SARS-CoV-2, the virus responsible for COVID-19, has undergone significant evolution since its emergence, leading to waves of outbreaks by multiple variants such as Alpha, Beta, Gamma, Delta, and Omicrons. Each of these variants exhibits distinct characteristics in terms of transmissibility, pathogenicity, and ability to evade immune responses. A key driver of this evolution is the mutation of the spike-protein-encoding gene, particularly in the receptor-binding domain (RBD), which is crucial for the virus’s ability to bind to host cell receptors and initiate infection^1–3^. Mutations in the RBD can enhance the virus’s ability to spread and escape immune detection^4^, making it a critical focus for variant tracking and public health monitoring. For example, the Delta variant harbors the L452R and T478K mutations which increases binding affinity to ACE2 receptor on host cells and contributes to reduced vaccine efficacy^5–9^. Understanding and tracking these variants are crucial for managing ongoing and future outbreaks.

Traditional surveillance methods, such as genome sequencing of clinically ascertained samples, have been instrumental in identifying and tracking SARS-CoV-2 variants. However, these methods are often time-consuming, expensive, and resource-intensive^10–12^. Moreover, clinical testing primarily captures data from symptomatic individuals, potentially missing large portions of the infected population who are asymptomatic or pre-symptomatic^13–16^. In contrast, wastewater surveillance has emerged as a promising approach to monitor community-level prevalence and diversity of SARS-CoV-2. By analyzing wastewater samples, one can detect viral RNA shed by infected individuals, regardless of their symptom status^17–19^. Wastewater surveillance offers several advantages, including non-invasiveness, cost-effectiveness, and the ability to capture data from a broad segment of the population. This method has now been widely implemented globally, providing early warnings of outbreaks and offering a comprehensive view of community-level viral transmission.

Wastewater sequencing is a crucial tool for early detection of viral variants. By collecting and analyzing the nucleic acids in wastewater using next-generation sequencing, researchers can identify mutations and new variants weeks before they appear in clinical samples^20–22^. This method allows continuous tracking of variant dynamics and provides a more complete picture of the viral landscape^23–26^. However, the complex nature of wastewater, containing a mixture of genetic material from various sources, complicates data analysis and interpretation, posing challenges in genome assembly, variant assignment/classification, and sequencing accuracy^27–32^. Typically, wastewater sequencing relies on tiled amplicon amplification to recover the whole genome due to low concentrations. A simpler approach targets the spike genes, particularly the RBD, with specific target areas varying among studies ^21,33–38^. While existing research has primarily focused on the detection and surveillance of variants, there has been limited exploration into the epidemiological dynamics and patterns of variant emergence and evolution within community wastewater.

In this study, we targeted the receptor-binding domain region and developed a streamlined bioinformatic pipeline to track the variant dynamics of SARS-CoV-2 over 17 months using wastewater samples from El Paso, a border city in west Texas, US, and Ciudad Juárez, Mexico. We compared our wastewater data with clinical genome sequencing results at local, state, and national levels to assess consistency and reliability. Additionally, we conducted epidemiological dynamic analyses to uncover patterns of variant evolution and emergence. By integrating RBD amplicon sequencing with wastewater surveillance, this research offers an innovative approach to understanding variant emergence and evolution, enriching the surveillance toolkit to anticipate and mitigate emerging threats in the post-pandemic era.

## 2. Materials and Methods

### 2.1 Wastewater sampling

Composite wastewater samples (24-hr) were collected weekly from March 14th, 2022, to August 8th, 2023, from three wastewater treatment plants (WWTPs) in the City of El Paso, Texas. The WWTPs A, B, and C serve 98,754, 125,462, and 128,003 residents, respectively. The daily total wastewater flow volume was measured and provided by El Paso Water. Samples were shipped overnight in ice boxes to the laboratory and processed on the day of receipt. The rest of the raw samples were stored at -80 °C.

### 2.2 Wastewater sample processing, RNA extraction, and RT-qPCR

Samples were processed based on previous methods^39–41^. Briefly, 50 mL wastewater samples were first centrifuged at 3000 g for 10 min at 4°C. The supernatant was collected and further filtered with MilliporeSigma™ Steriflip™ Sterile Disposable Vacuum Filter Units. Then, the 15 mL filtrates were concentrated to ∼200uL with MilliporeSigma™ Amicon™ Ultra-15 Centrifugal Filter Units. The enriched samples were subject to RNA extraction with QIAamp Viral RNA Mini Kit according to the manufacturer’s protocol. RNA was eluted with 100 uL nuclease-free water and stored at -20°C.

SARS-CoV-2 was tested and quantified using the US CDC N1 real-time PCR assay (Biorad, CFX96 C1000 Touch) using the following program: 50 °C 10 min for reverse transcription, 95 °C 20 s for RT inactivation and initial denaturation, and 48 cycles of denature (95 °C, 1 s) and anneal/extend (60 °C, 30 s). At least two negative controls were included in every PCR run. Pepper mild mottle virus (PMMoV) was also measured as an internal reference for sample processing and variations in wastewater flow and/or fecal materials, as previously reported ^40,42^. Two technical replicates were performed for all RT-qPCR reactions. SARS-CoV-2 RNA concentrations were adjusted by the corresponding PMMoV concentrations in the sample using the method described in previous work^40,42^. The total viral load per capita was calculated for data comparison across sewersheds with the following formula:

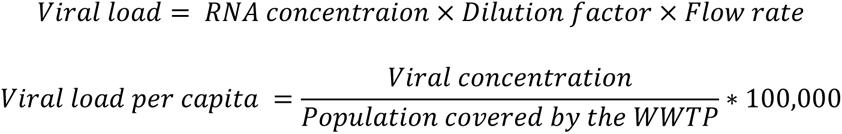

The Dilution Factor is the scale of enrichment during the experiment, which is 6.7 (100 uL/15 mL). This indicates that 15 mL of raw composite wastewater sample was concentrated into 100 uL of RNA.

### 2.3 Preparation of synthetic SARS-CoV-2 variant communities

The SARS-CoV-2 Wuhan-Hu strain and Omicron BA.4&5 genomes were purchased from TWIST Bioscience. Original stocks were diluted one thousand-fold before mixing with four different ratios: 10%:90%, 25%:75%, 50%:50%, and 90%:10%. 10 ul of each mixture sample were then subjected to RNA extraction with QIAamp Viral RNA Mini Kit.

### 2.4 RBD amplicon amplification of wastewatwer samples

Primers for amplifying the receptor-binding domain (RBD) of the SARS-CoV-2 spike glycoprotein were provided in Table S1 and adapted from a previous study^20^. This amplicon amplifies 168 amino acids from 337 to 504 aa, covering almost the entire receptor-binding motif of SARS-CoV-2 (from 437 to 508 aa) and most receptor-binding domain (from 319 to 541 aa). Briefly, wastewater samples and positive control RNA were transcribed into cDNA using SuperScript™ IV Reverse Transcriptase (ThermoFisher Scientific, USA). The RBD amplicon was then amplified from the cDNA using primers S008 and S012 in a first PCR with Q5® High-Fidelity 2X Master Mix (New England Biolabs, USA), under the following conditions: polymerase activation at 95°C for 2 minutes, followed by 30 cycles of denaturation at 95°C for 1 second, annealing at 72°C for 30 seconds, and extension at 60°C for 30 seconds. A second PCR was performed to add Illumina overhangs to the RBD amplicon using primers S009 and S013, with the same conditions but for 40 cycles. The PCR product was purified using AMPure XP beads and eluted with 30 µL of nuclease-free water. For testing the protocol, synthetic SARS-CoV-2 Wuhan-Hu strain and Omicron BA.4&5 genomes from TWIST Bioscience were diluted 1,000-fold, and four mixtures were prepared in ratios of 10%:90%, 25%:75%, 50%:50%, and 90%:10%. 10 µL of each mixture was used for RNA extraction, amplicon amplification, library preparation, and sequencing.

### 2.5 Library preparation and sequencing data analysis

The purified PCR amplicons were dual-indexed using NEBNext Multiplex Oligos (New England Biolabs, USA) for Illumina and further purified using AMPure XP beads. Libraries were pooled based on their concentrations and sequenced on a 300 paired-end Miseq run at the ATGC Facility at MD Anderson Cancer Center. The raw sequencing data in fastq format were de-multiplexed. DADA2 package (version 4.3.0)^43^ was used to filter and trim sequences, merge paired ends, and remove chimeras in Rstudio version 4.3.2. The unique amplicon sequence variants (ASVs) were aligned and identified using the BLAST online tool^44^ using the Betacoronavirus database. For each ASV, the top 10 BLAST results containing the Pango Lineage name with the highest alignment scores (ranging from 99% to 100%) were downloaded, and the most frequent variant was identified as the variant for that sequence. The relative abundance of each variant was calculated by the ratio of the count of each variant to the total sequence count. Variants with relative abundance higher than 10% on at least one sample were designated as major variants. The remaining variants were grouped as ’others’ for plotting and visualization purposes.

### 2.6 COVID-19 cases and genome data

The reported COVID-19 case data by lab-testing diagnostics was downloaded from healthdata.gov, and the 7-day rolling average was used for further analysis and data visualization. The Texas-specific data was extracted according to the state name label. SARS-CoV-2 genome data for the study period was obtained from the GISAID EpiCoV^TM^ database using the GISAIDR (version 0.9.9) package in R^45^. The variants were assigned by Pango v.4.3 by GISAID. Only records with complete genomes were downloaded. The number of each variant per week was counted, and the relative abundance was calculated accordingly.

## 3. Results

### 3.1 RBD amplicon sequencing of wastewater reveals temporal dynamics of SARS-CoV-2 variants circulating in the city

To validate the RBD amplicon sequencing approach for variant identification, we utilized synthetic mock variant communities. Four mock communities were prepared by mixing the original SARS-CoV-2 Wuhan-Hu strain (Wild Type) and the BA.4&5 strain at ratios of 10%:90%, 25%:75, 50%:50%, and 90%:10%. Each mock community underwent RNA extraction, reverse transcription, amplicon amplification, library preparation, and sequencing, followed by bioinformatic analysis. As a negative control, nuclease-free water was processed identically, except for RNA extraction. The sequencing results showed that nearly all reads (98.5%∼100%) were accurately identified as either Wild Type or Omicron BA.4&5 strains across all four mock communities. The estimated relative abundances for each variant in the mock communities were 6.7%:92.3%, 21.9%:76.6%, 51.3%:47.5%, and 93.1%:6.9% (**Figure 1B**), closely matching the original mixing ratios with a deviation range of -3.1% to 3.1%. These results demonstrate that the RBD amplicon sequencing method is effective for identifying variants and estimating the composition and structure of SARS-CoV-2 variants in a sample.

**Figure 1.**
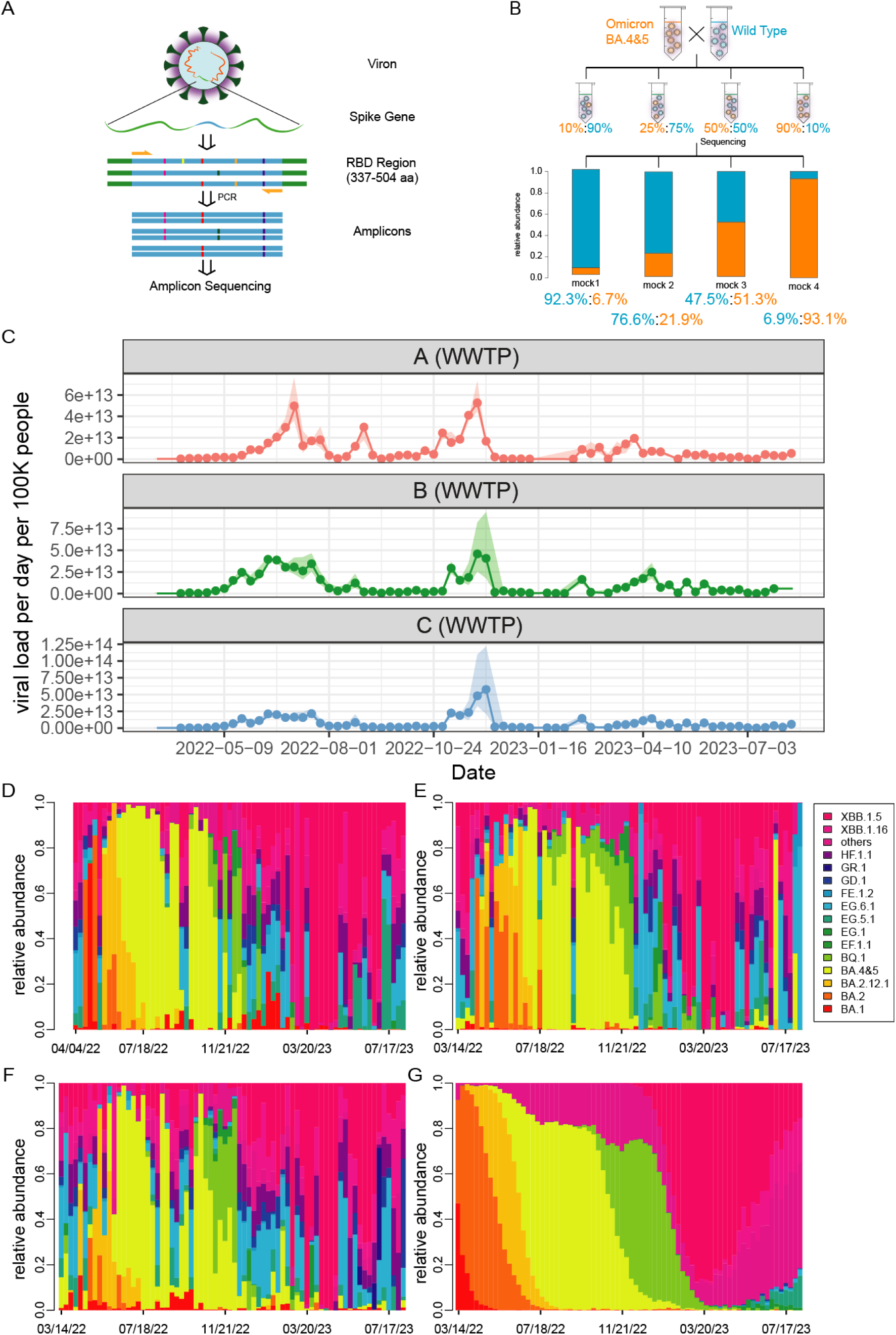
RBD amplicon sequencing of wastewater reveals temporal dynamics of SARS-CoV-2 variants. (A) Diagram of the RBD amplicon sequencing procedure. We specifically amplify the 337-504 aa sequence in the RBD region of SARS-CoV-2 spike gene. (B) Validation of the RBD amplicon sequencing protocol using the wild-type SARS-CoV-2 strain and BA.4&5. The bar plot shows the sequencing results of these samples, demonstrating that the resultant ratios closely match the original mixing ratios. (C) Viral load of SARS-CoV-2 per day per 100k people in the three sewersheds in the City of El Paso, TX from March 2022 to August 2023. (D-F) Relative abundance of SARS-CoV-2 variants revealed by RBD amplicon sequencing of wastewater samples from WWTPs A (D), B (E), and C (F) in the City of El Paso. (G) Relative abundance of SARS-CoV-2 variants from genomic sequencing of clinical samples in Texas.

Next, we applied the RBD amplicon sequencing approach to weekly wastewater samples collected from the three wastewater treatment plants. All samples tested positive for SARS-CoV-2. Consistent with clinically reported case data, viral load results revealed three major waves of infection in the city: from April 2022 to September 2022, from November 2022 to December 2022, and from February 2023 to the end of the experiment period (August 2023) (**Figure 1C and Figure S1**).

Across the 216 samples analyzed, 91 SARS-CoV-2 variants were identified, each with over 99% identity to the reported variant sequences. For visualization, we plotted the related abundance of the variants of interest (VOIs) listed by WHO and those with an abundance of 10% or higher in at least one wastewater sample. All other variants were grouped under ’others’. **Figures 1D-F** show the relative abundance of SARS-CoV-2 variants over the 17 months across the three sewersheds, highlighting the evolving composition of variant communities. Overall, each wastewater sample contained multiple variants, and we observed waves of dominant variants transitioning from BA.2 to BA.2.12.1, BA.4&5, BQ.1, and to XBB.1.5. While there were some differences, the overall variant profiles for each WWTP were similar, indicating that the spread and evolution of the virus followed a comparable trajectory across the three sewersheds in the city. These results demonstrate that RBD amplicon sequencing of wastewater samples can be used to monitor the temporal dynamics and community structure of SARS-CoV-2 variants circulating in the population.

### 3.2 Consistency and novel insights from RBD amplicon sequencing data compared to clinical genome sequencing data

We further validated the wastewater findings using reported clinical genome sequencing data from the USA, Texas, and El Paso. As shown in **Figure 1G**, the overall trends between the wastewater RBD amplicon sequencing data and clinical genome sequence data in Texas are consistent, with multiple waves of major variants emerging sequentially. Moreover, the composition of variants in wastewater samples is more diverse, suggesting that some variants circulating in the city may go undetected in clinical testing. We further compared the temporal dynamics of major variants during the study period: BA.2, BA.2.12.1, BA.4&5, BQ.1, and XBB.1.5 (**Figure 2**) in both wastewater and clinical sequencing data at the El Paso, Texas, and national levels. All five variant waves were consistently found in both datasets, showing similar transmission waves and relative abundance at the peaks. We also noticed some differences between Texas and national trends. For example, the BA.2 peak occurred later, and the BA.4&5 peak arrived earlier and lasted longer in Texas compared to the national data. However, wastewater data in El Paso closely matched the clinical sequencing data in Texas.

**Figure 2.**
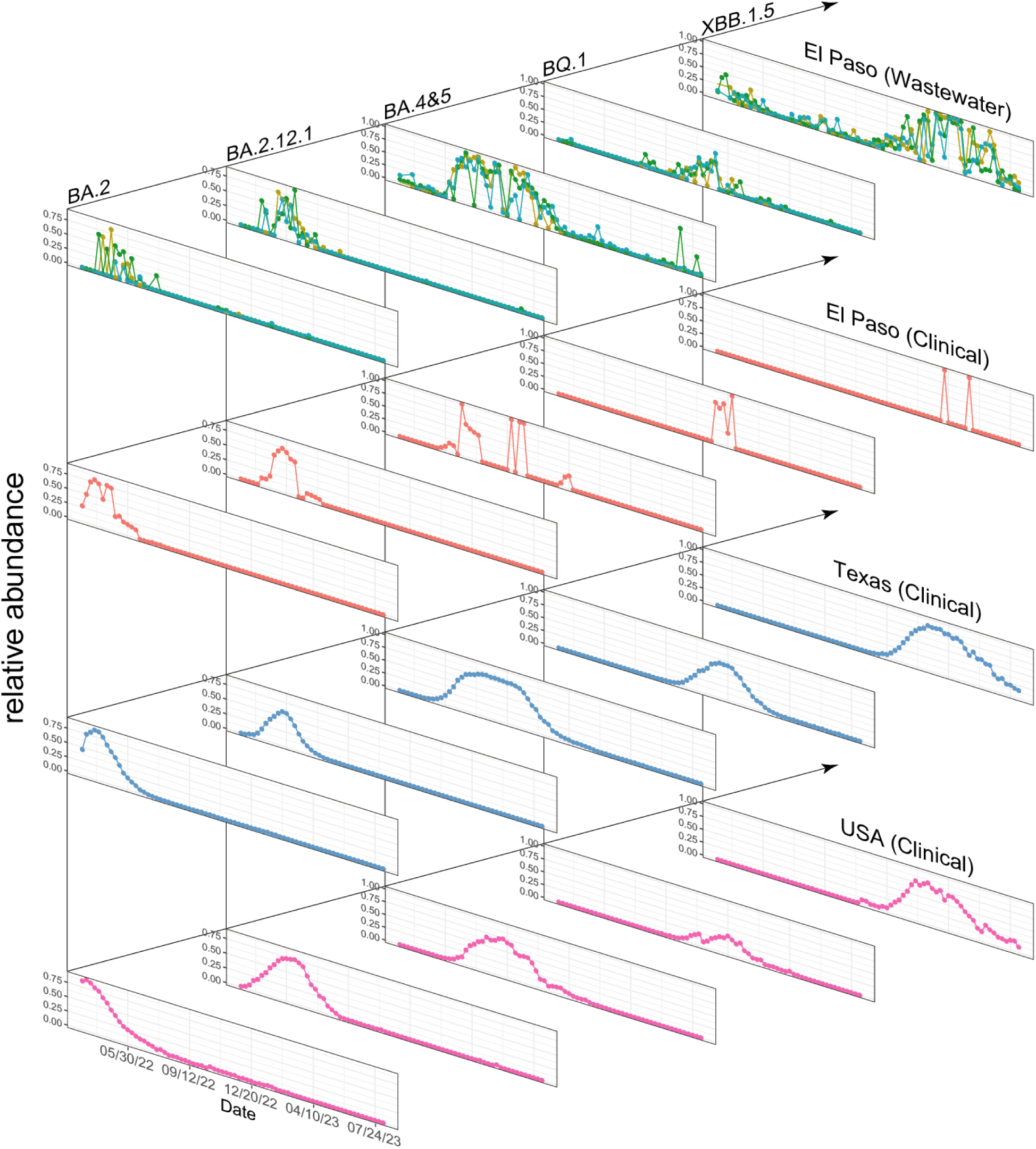
Time-course of major SARS-CoV-2 variants identified from RBD amplicon sequencing of wastewater. This figure presents the temporal dynamics of five major variants identified from three WWTPs alongside clinical data from El Paso, Texas, and the USA. Rows depict the progression of variant waves over time, while columns allow for a comparative analysis between the wastewater treatment plant data and clinical data, highlighting their similarities and trends.

In addition, we found that some variants circulated in the population earlier than their prevalence observed in clinical tests. For example, the XBB.1.5 wave began in December 2022, as shown in both clinical and wastewater data (Figure 2). However, this variant was detectable in wastewater samples before the major wave, with small peaks observed in March and September 2022 (**Figure 2**). Similarly, the waves of EG.5.1 and XBB.1.16 started in May and March 2023, respectively (**Figure 3A and 3B**), yet these variants were detectable from the beginning of our experiment. These findings strengthen the existing idea that wastewater surveillance could provide early warnings to the community ^28,46,47^.

**Figure 3.**
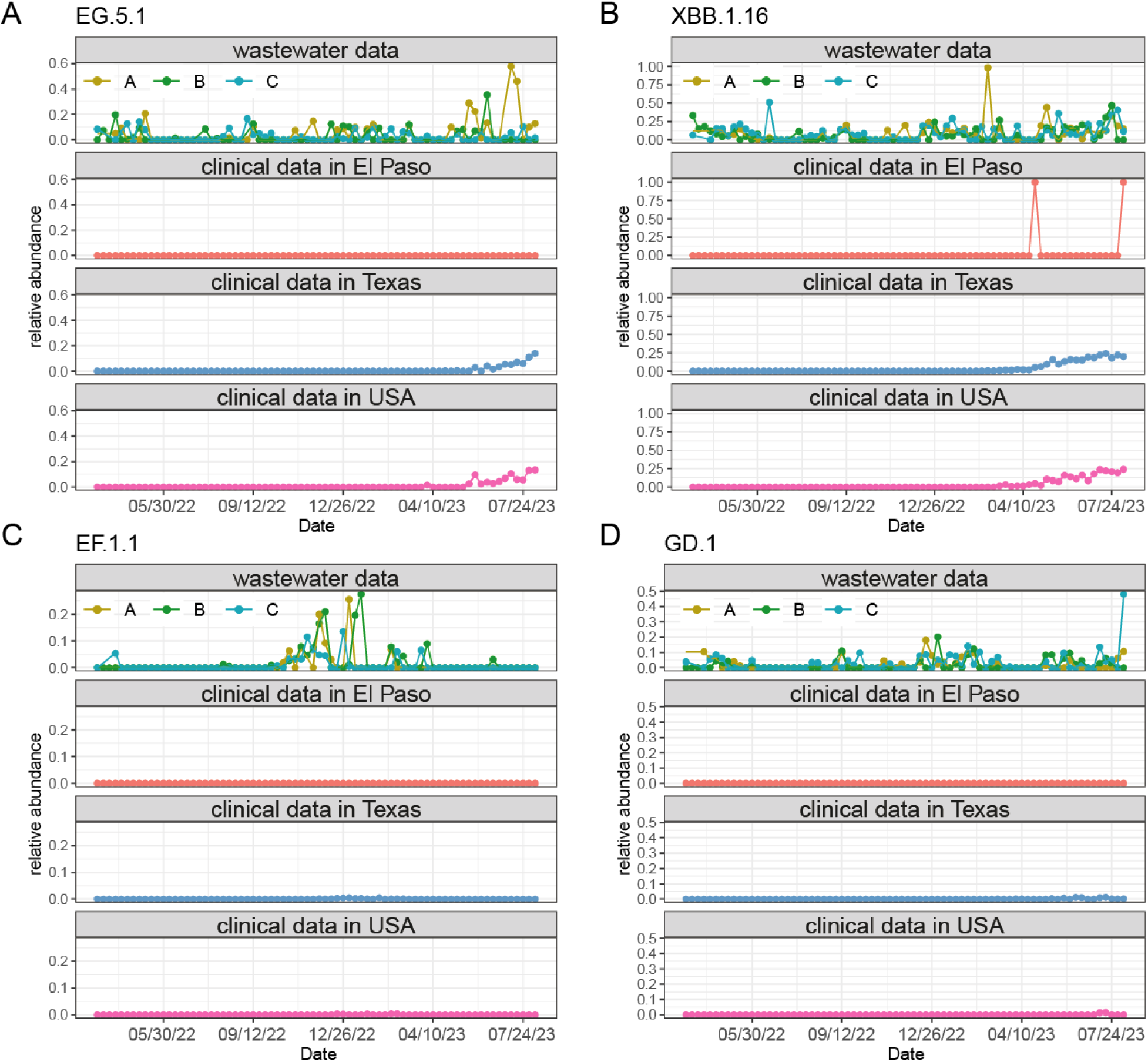
Early detection and identification of unreported SARS-CoV-2 variants from RBD amplicon sequencing data. (A-B) The EG.5.1 and XBB.1.16 variants were detected circulating in the city earlier than their corresponding waves in clinical genome sequencing data from El Paso, Texas, and the USA. (C-D) The EF.1.1 and GD.1 variants were found in the city but were rarely reported in clinical genome sequencing data.

We also identified some uncommon variants that were not detected in the clinical data. Those variants, including EF.1.1, GD.1, EG.6.1, and FE.1.2, were barely noticeable in clinical records due to low abundance but were significant enough to exceed 10% in at least one wastewater sample (**Figure 3C and 3D, and Figure S2**). The EF.1.1 variant even had a small outbreak in all of the three sewersheds, with a relative abundance of up to 25% from October 2022 to February 2023. The EG.6.1, FE.1.2, and GD.1 variants were detected throughout the experiment period, but very few records were found in clinical data. This discrepancy likely occurs because these variants have a moderate transmission rate but do not cause severe symptoms, leading to fewer clinical tests.

In summary, the results from RBD amplicon sequencing of wastewater samples are consistent with clinical genome sequencing data in capturing the variants’ dynamics. This consistency highlights the effectiveness and accuracy of this approach in monitoring SARS-CoV-2 variants circulating in the sewershed. Additionally, the method allows for the early detection of emerging variants and provides a comprehensive view of variant diversity, emphasizing its importance for community-level surveillance efforts.

### 3.3 RBD amplicon sequencing data highlights off-season and lag-phase as crucial periods for variant emergency and evolution

To better understand the epidemiological dynamics of SARS-CoV-2, we performed alpha diversity analyses of the variants identified from the RBD amplicon sequencing of wastewater samples. Specifically, we examined how variant diversity changes with the seasons, during high transmission periods (in outbreak waves) versus low transmission periods (out of wave), and across different outbreak phases including lag, growth, stationary, and decline. Metrics including Observed, Chao1, Shannon, and Inverse Simpson indices were used to measure the richness and evenness of SARS-CoV-2 variants in the wastewater samples. The Observed index counts the actual number of unique variants, Chao1 estimates total species richness, Shannon quantifies the diversity considering both abundance and evenness, and Inverse Simpson emphasizes the dominance of the most abundant variants.

Results revealed significant seasonal variations in alpha diversity in all four metrics (**Figure 4A**). An increasing trend of alpha diversity was observed from spring to winter, with the highest alpha diversity observed in winter, suggesting that many variants emerged during the winter months. And summer months exhibited the lowest diversity. Moreover, higher diversity was also observed during the low transmission (out of wave) period across the four metrics (**Figure 4B and 4C**). This suggests that during these low transmission periods, a more diverse pool of variants circulates at low levels in the population, potentially due to reduced selective pressures compared to the outbreak period. When grouping data into different outbreak phases, we found that the highest diversity emerged in the lag phase, followed by the decline and growth phases, with the lowest diversity observed in the stationary phase (**Figure 4D and 4E**). This result suggests an interesting phenomenon: new variants mostly emerged not during the stationary phase, where the highest viral shedding and new cases occur, but in the lag phase, before a new outbreak starts. Beta diversity analysis showed no significant difference across the three sewersheds (**Figure 4F**). Together, these results showed that RBD amplicon sequencing of wastewater is a powerful tool for uncovering the patterns of variant emergence and evolution in the sewershed.

**Figure 4.**
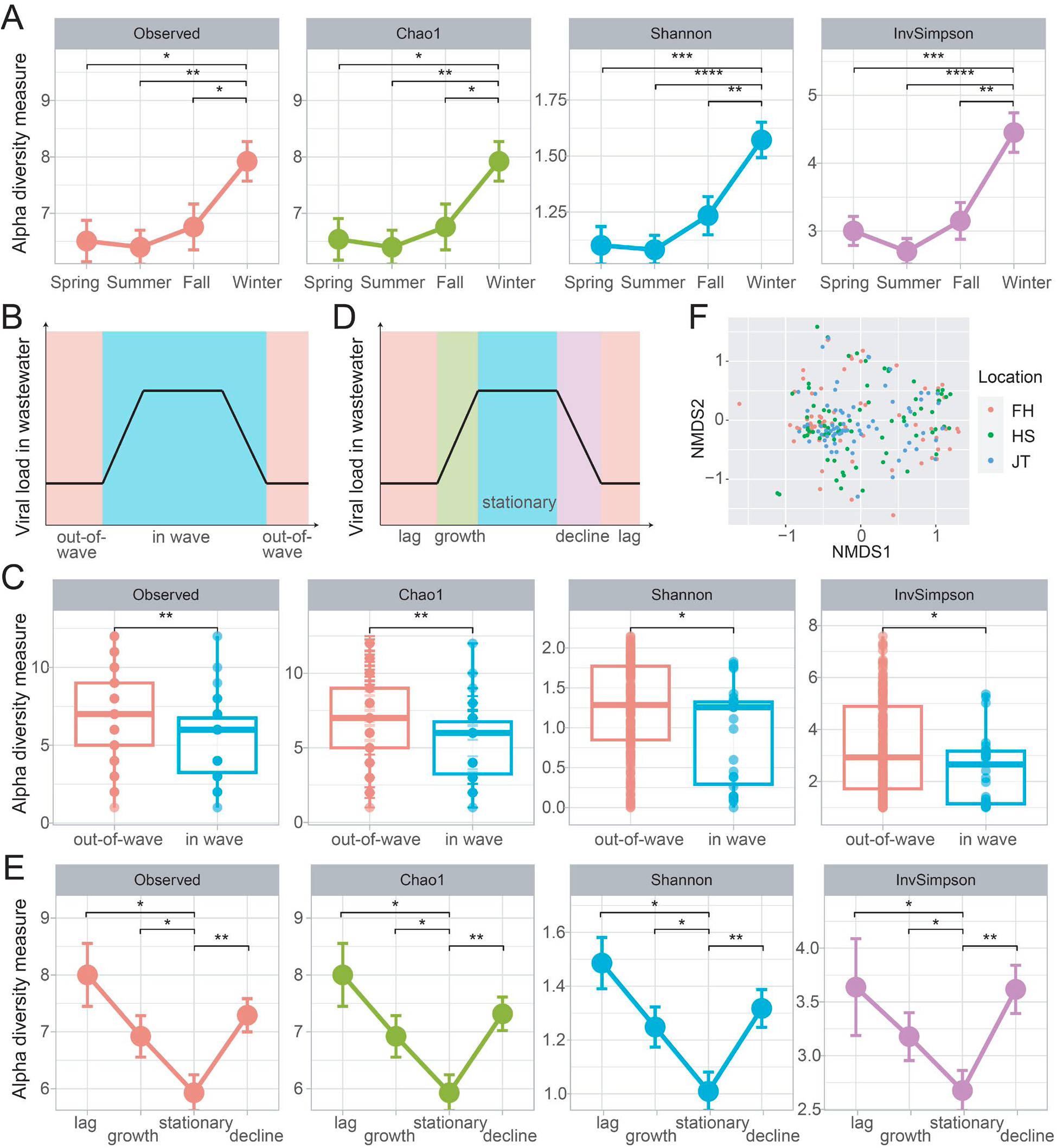
Epidemiological patterns of variant emergence and evolution. (A) Alpha diversity comparison across seasons in the three sewersheds. (B) Schematic diagram of the high transmission (in-wave) and low transmission periods (out-of-wave) in an epidemic cycle. (C) Comparison of alpha diversity between in-wave and out-of-wave samples. (D) Schematic diagram of an epidemic cycle: lag, growth, stationary, and decline. (E) Alpha diversity comparison across the four phases: lag, growth, stationary, and decline. (F) Beta diversity analysis across all three WWTPs. No significant differences in viral diversity among the three sewersheds. Error bar: standard error; Wilcoxon rank-sum test was used for the group comparisons with significance: *: p< 0.05; **: p<0.01; ***p<0.001. ****: p<0.0001.

## Discussion

One significant insight from this study is the ability of RBD amplicon sequencing data to reveal patterns of viral evolution and variant emergence. Previous studies using whole genome sequencing of SARS-CoV-2 in wastewater have highlighted the early-warning potential of this approach and its ability to uncover variant dynamics^48–51^. However, our study emphasizes that wastewater data can also be viewed from an ecological and evolutionary perspective, reflecting the community’s viral variant landscape.

Our seasonal analysis revealed significant variations in alpha diversity, with higher diversity observed in winter and lower diversity in summer. This suggests that colder months facilitate the emergence and spread of more variants, likely due to increased indoor activities, enhanced viral stability, and seasonal variations in immune responses. Additionally, higher alpha diversity during low transmission periods and the lag phase of outbreaks indicates that a diverse pool of variants circulates at low levels during these times. These periods allow for the accumulation and persistence of a wider range of variants, which may not immediately lead to outbreaks but could contribute to future waves if conditions become favorable. Ecologically, the lag phase represents a period of low competition, allowing new variants to compete until a strain with the highest fitness prevails. These insights underscore the importance of continuous monitoring of viral diversity to predict future outbreaks and implement timely public health interventions.

Our findings also demonstrate that RBD amplicon sequencing of wastewater samples effectively captures the dynamics of SARS-CoV-2 variants, showing consistent results with clinical genome sequencing data at local, state, and national levels. Notably, our study identified variants circulating in the population earlier than observed in clinical sequencing data and detected variants and outbreaks that were not reported or noticed locally. This consistency with clinical genome sequencing data and the earlier detection of variants underscore the accuracy and reliability of RBD amplicon sequencing of wastewater in variant surveillance. It also strengthens the role of wastewater surveillance as a complementary tool to clinical testing^24–26,52–55^, especially valuable in areas with limited access to clinical sequencing resources.

Despite its advantages, RBD amplicon sequencing has limitations. While the RBD region is rich in information and useful for targeted analysis, it represents only a small portion of the viral genome. This limitation means that mutations outside the RBD region will be missed, and closely related variants with mutations in other regions may not be differentiated. Additionally, evolutionary changes occurring outside the targeted region, including recombination events, are not captured. The PCR amplification step can also introduce biases^56–58^, potentially skewing the relative abundance of certain variants. While our analyses using synthetic variant communities (**Figure 1B**) suggest that this bias may not significantly impact our results, further validation is necessary. On the other hand, RBD amplicon sequencing is faster, easier, and cheaper than whole genome sequencing^20,59,60^, making it accessible for widespread use. Its high throughput capability with relatively lower costs allows for the rapid analysis of numerous samples, which is critical during pandemic conditions. By balancing these trade-offs, RBD amplicon sequencing provides a practical and efficient tool for ongoing variant surveillance, particularly when rapid data collection is necessary and resources are constrained.

Viruses have various mechanisms to enhance transmission between hosts. One significant strategy is mutating the receptor-binding domain in the spike gene, as observed with SARS-CoV-2, which has produced multiple waves of variants from Alpha to Omicron. These mutations in RBD enable the virus to evade adaptive immune responses from prior infections and/or vaccinations. Consequently, monitoring the RBD region is crucial for tracking viral evolution, predicting the emergence of new variants, and informing updated vaccine composition. This study highlights the utility of RBD amplicon sequencing and the integration of wastewater surveillance to track SARS-CoV-2 evolution and variant emergence. Our findings demonstrate that this tool captures the dynamics of variant community, provides early detection of variants, aligns well with clinical genome sequencing data, and reveals epidemiological patterns in viral diversity. The concept of monitoring the receptor-binding domain can also be generalized to other viruses, providing a broader framework for variant surveillance. This tool can improve our ability to predict future outbreaks and enhance public health preparedness.

## Data Availability

All data produced in the present study are available upon reasonable request to the authors

## Data and Code Availability

Data in this study will be shared with the paper publication. Scripts will be shared on GitHub.

## Acknowledgments

We are grateful to Teresa T. Alcala at El Paso Water for sample collection and shipping. This work is supported by the PRIME award from the UTHealth Houston School of Public Health, National Institute of Health RADxUP (1U01TR004355-01), and the Texas Epidemic Public Health Institute (TEPHI).

