## Supplementary Figure for "RBD amplicon sequencing of wastewater reveals patterns of variant emergence and evolution"

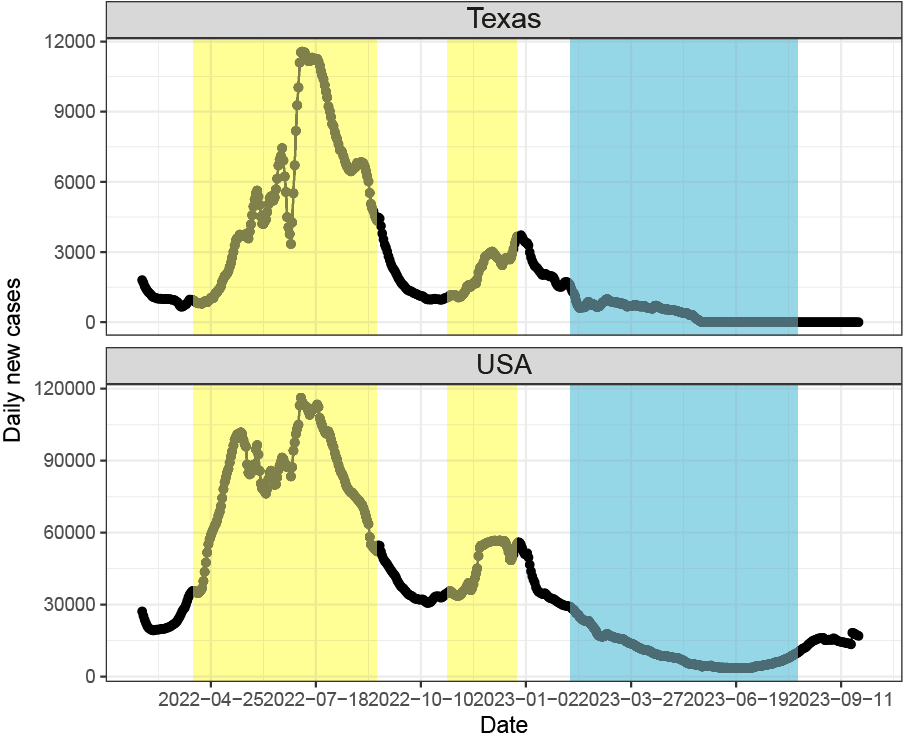

**Supplementary Figure 1. Daily new cases over time of Texas and the USA.** The yellow shaded period shows the two waves identified by both clinical data and our wastewater data. The blue shaded period shows the wave identified only in wastewater data.

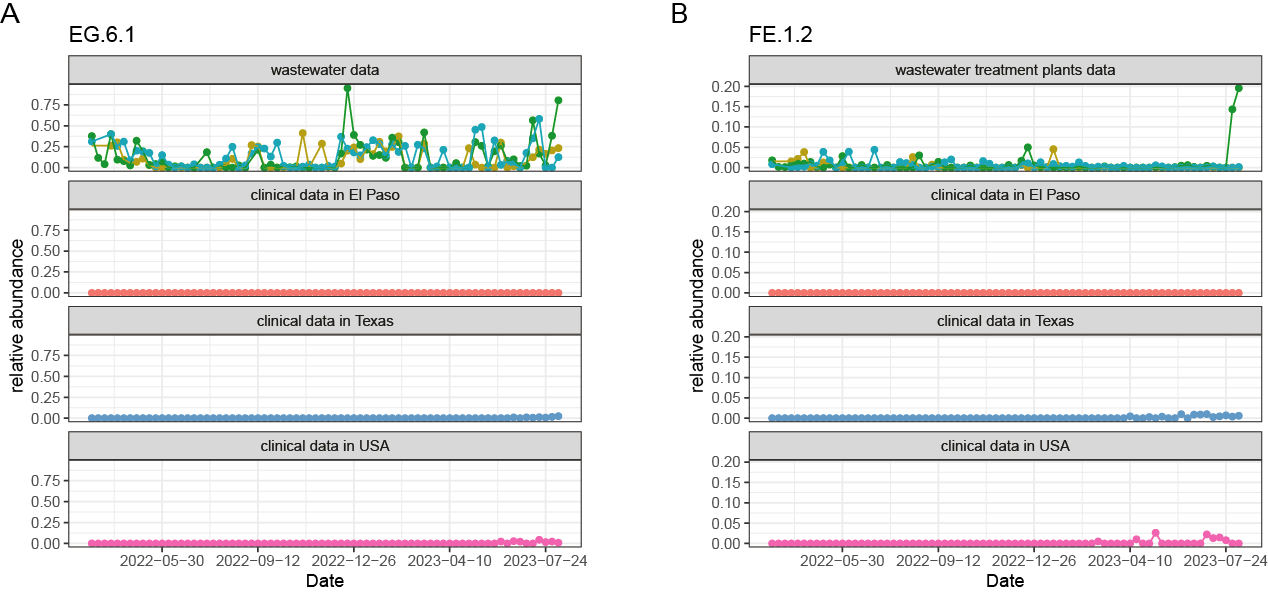

**Supplementary Figure 2. Extra** **examples of variants with wastewater and clinical data discrepancies.** The EG.6.1 and FE.1.2 variants were found in the city but were rarely reported in clinical data during this study.

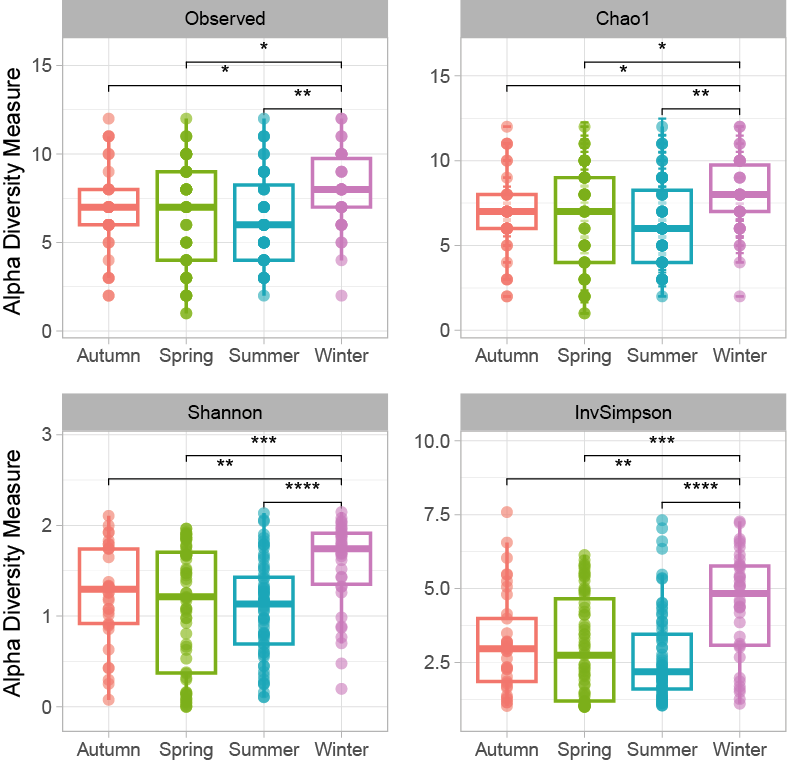

**Supplementary Figure 4. Alpha diversity of seasons.** Wilcoxon rank-sum test was used for the group comparisons with significance: *: p< 0.05; **: p<0.01; ***p<0.001.

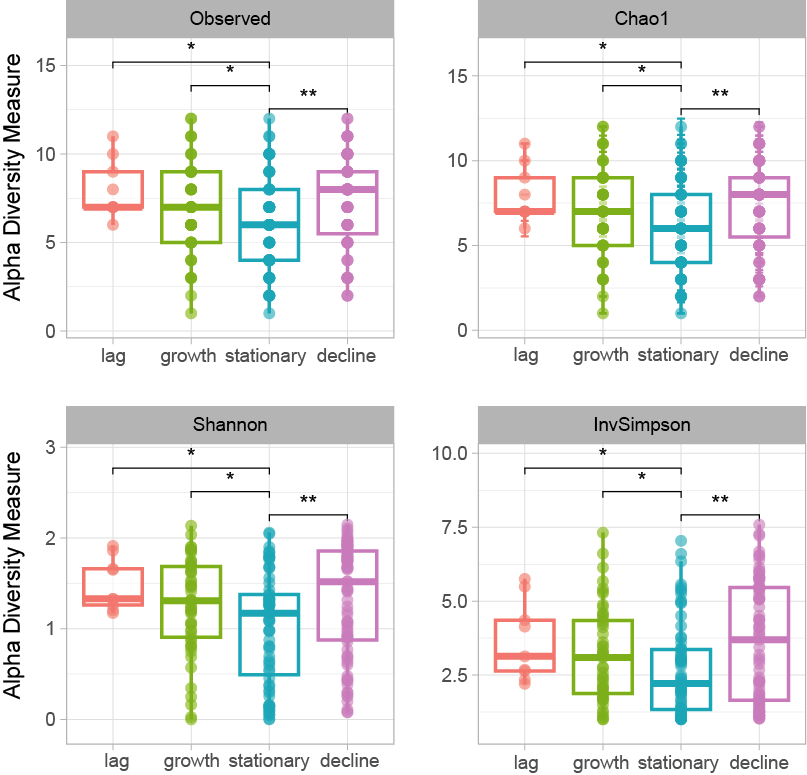

**Supplementary Figure 4. Alpha diversity of different phases.** Wilcoxon rank-sum test was used for the group comparisons with significance: *: p< 0.05; **: p<0.01; ***p<0.001.
